# Modeling the impact of outbreak response immunization on typhoid fever using data from 19 outbreaks between 2000-2022

**DOI:** 10.64898/2026.04.28.26351891

**Authors:** Monica Duong, Jong-Hoon Kim

**Affiliations:** Epidemiology, Public Health, Impact, International Vaccine Institute, Seoul, South Korea

**Keywords:** Typhoid, Typhoid conjugate vaccine, Outbreak response immunization, Vaccine impact modeling

## Abstract

Typhoid fever remains endemic in many low- and middle-income countries, with antimicrobial-resistant *S*. Typhi increasingly triggering outbreaks in previously unaffected regions. While typhoid conjugate vaccines (TCVs) offer a means of outbreak control, the potential impact of outbreak response immunization (ORI) — and how coverage, timing, and outbreak characteristics shape that impact — remains poorly characterized. We evaluated single-dose TCV ORI strategies using a time series dataset of 19 typhoid outbreaks reported between 2000 and 2022. A static model was applied to each outbreak, incorporating direct and indirect vaccine effectiveness, immunological delay, and cost-effectiveness analyses across a range of timing and coverage scenarios. The 19 outbreaks had a median duration of 27.5 weeks and median size of 805.5 cases. At an 8-week ORI delay and 80% vaccine coverage, outbreaks reporting weekly incidence showed a median case reduction of 30.0% (95%CI: 0%–68.5%), corresponding to a median of 275 cases averted (95%CI: 0–2,787). The median cost per DALY averted was $82,756 (95%CI: 0–618,927) and median cases averted per 1,000 doses was 0.63 (95%CI: 0.06–8.76). ORI timing, vaccine coverage, and outbreak characteristics — particularly duration and size — were key determinants of impact. ORI is a valuable strategy for mitigating typhoid fever outbreaks. Timely deployment substantially reduces case burden, particularly in larger or longer outbreaks, and is most effective when integrated with water, sanitation, and hygiene interventions in resource-limited settings.

## Introduction

Typhoid fever is a systemic illness caused by the Gram-negative bacterium *Salmonella enterica* serovar Typhi (*S. Typhi*), and it remains endemic in many low- and middle-income countries, particularly where clean water and adequate sanitation are lacking.^13^ In 2021, an estimated 7.15 million cases of typhoid fever occurred globally (95% CI: 5.57 – 9.23 million), resulting in approximately 93,000 deaths (95% CI: 46,257 – 158,181)^14^. Although effective antibiotics can reduce typhoid case fatality rates to below 1%, the growing emergence of antimicrobial-resistant *S. Typhi* poses a serious public health threat, sparking outbreaks in regions that had not previously been affected^15^. Between 2000 and 2022, 43 typhoid outbreaks were reported in the literature^16^.

Typhoid conjugate vaccines (TCV) offer an effective method for prevention and mitigation of outbreaks, especially as a short-term tool in places that require time and investments into improving access to clean water, sanitation, and hygiene infrastructure^17^. Since 2018 and 2020, two single-dose TCVs have been available, Typbar TCV® (Bharat Biotech) and TYPHIBEV® (Biological E), while two more TCVs were recently prequalified by the World Health Organization (WHO) in 2024, SKYTyphoid™ (SK bioscience) and ZyVac® TCV (Zydus Lifesciences Limited)^18–20^. The WHO recommends TCV for routine administration as a single dose to children from six months of age to adults of 45 years of age living in endemic regions, as well as for use in catch-up campaigns for those up to 15 years old^19^.

Since 2017, five countries have received Gavi support for routine immunization and catch-up campaigns using TCVs. However, only Zimbabwe has been granted funding specifically for outbreak response in 2019, despite the occurrence of typhoid outbreaks in other countries^19^. This limited response may stem from challenges in reporting, confirming, and characterizing the local epidemiology of typhoid, which hinders the accurate estimation of TCV needs. According to UNICEF (2022), while an annual supply of 6.5 million TCV doses is considered secure in the coming years, actual demand may range between 42.7 and 77.1 million doses^19^. Mathematical modeling offers a valuable tool for understanding typhoid transmission dynamics and evaluating the potential impact of TCVs. However, most prior modeling efforts have centered on routine or preventive vaccination in endemic settings, often limited to single or few locations. As epidemic outbreaks become increasingly concerning, partly due to the rise of antimicrobial-resistant *S. Typhi*, there is a pressing need to study the impact of outbreak response immunization (ORI) strategies to inform future TCV deployment and policy decisions.

This study utilizes a dataset of 43 typhoid outbreaks reported between 2000 and 2022 compiled by Koh et al. (2025), to assess the potential impact of ORI in outbreaks that range in size, duration, and geographic and temporal contexts. Our analysis incorporates key parameters including direct and indirect vaccine effectiveness, delays in vaccine-derived protection, as well as costs related to both vaccination and illness. Using these inputs, we estimate measures of vaccine impact and cost-effectiveness such as percent case reduction, cases averted per 1,000 doses, and costs per DALY averted.

## Methods

A total of 43 typhoid outbreaks between 2000 and 2022 were reported in the dataset by Koh et al. (2025). Of these, twenty-four outbreaks were excluded for being in a Gavi funding-ineligible country (as of 2025)^21^ (n = 17), having a prior vaccination campaign (n = 2), being very short (n = 1), and having other nested location-specific outbreaks already in the dataset (n = 2). A final total of 19 outbreaks were included for modeling, in which 6 reported daily incidence of suspected cases, 7 reported weekly incidence, 1 reported fortnightly incidence, and 5 reported monthly incidence (Figure 1).

**Figure 1.**
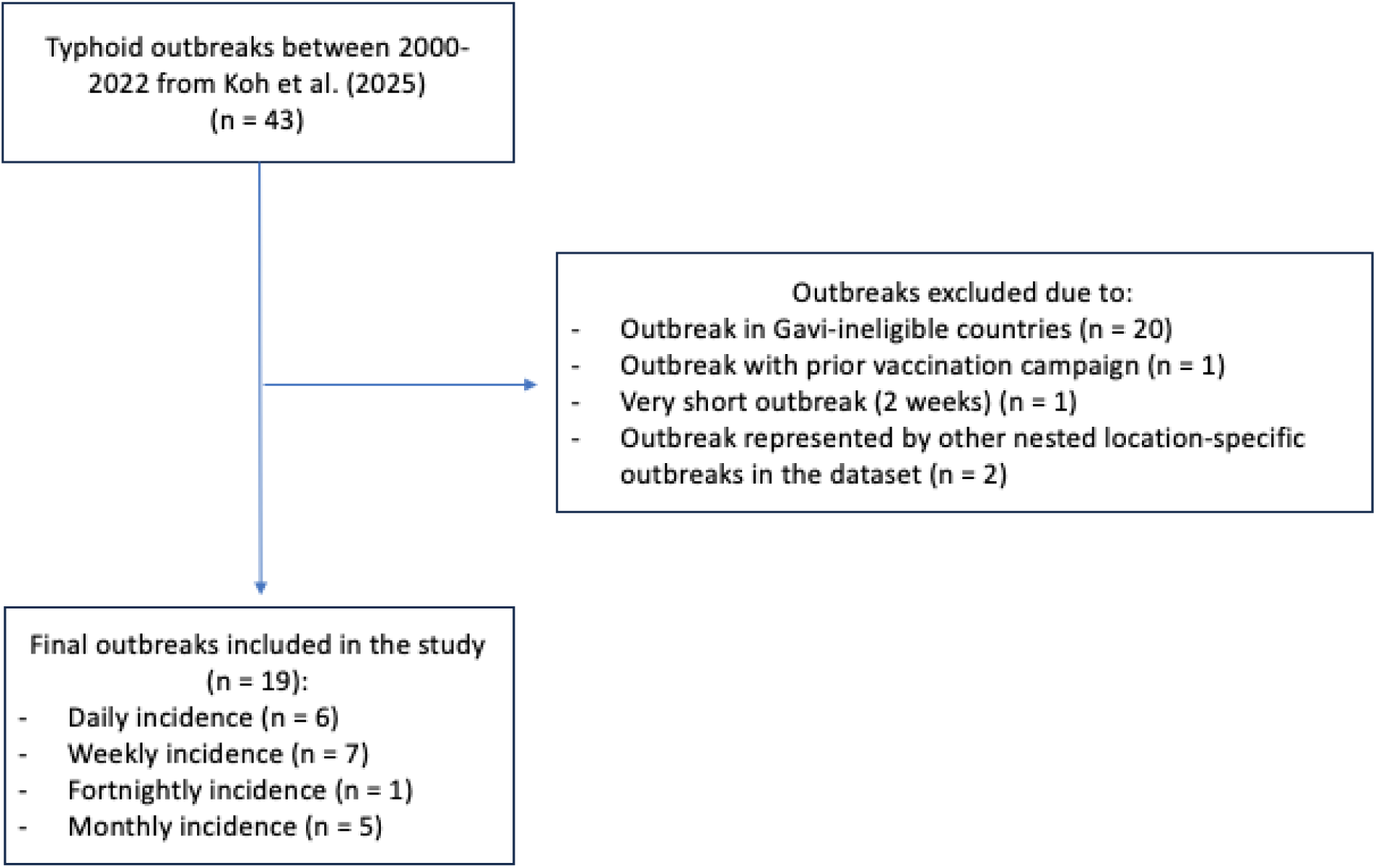
Overview of final typhoid outbreaks included in the study.

A static model was employed to model the impact of TCV on outbreaks. Since the focus is on epidemic outbreaks in native settings, the model focuses on the immediate impact of ORI and does not consider recurrent campaigns in the same population. The overall vaccine effectiveness, *P*_*i*,*π*_, is estimated using the following equation^22^:

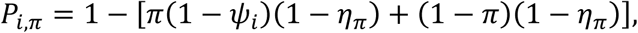

Where *P*_*i*,*π*_ represents the proportion of cases averted for age group *i* and vaccine coverage *π*, *η*_*π*_ represents the indirect vaccine effectiveness, and *ψ*_*i*_ represents the direct vaccine effectiveness.

The cumulative number of cases under a given vaccination campaign is then calculated as follows:

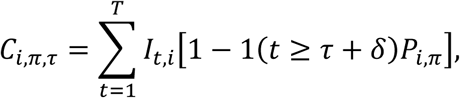

Where *C*_*i*,*π*,*τ*_ represents the cumulative number of cases for age group *i*, vaccine coverage *π*, and time *τ*, *I*_*t*,*i*_represents the incidence at time *t* for age group *i*, and 1(*x*) is an indicator function that equals 1 if *x* is true and 0 otherwise. The term *δ* represents the delays in campaign implementation and development of vaccine-derived immunity, and *T* denotes the outbreak’s end date.

Each outbreak was analyzed individually and the models compared counterfactual case reduction from vaccination scenarios with observed outbreaks in which no vaccination occurs. Outbreaks that reported daily incidence were aggregated to the weekly scale and the outbreak that reported fortnightly incidence was aggregated to the monthly scale. To align with previously published studies, the main text will present results for outbreaks on the weekly time unit. Additional analyses for outbreaks on the monthly time unit are also performed and presented in the supplementary material.

Figure 2 presents hypothetical counterfactual outbreak scenarios, highlighting the potential range of vaccine impact across a diverse set of outbreaks. For example, some outbreaks may conclude before an ORI can be initiated (blue), while others, despite lasting longer, may still be too brief to observe any measurable effect (orange) due to delays in the onset of vaccine-induced immunity. Impact is also influenced by outbreak size, where larger outbreaks could show greater impact (green) compared to outbreaks with smaller weekly cases (purple). However, timing remains critical because if an ORI is initiated too late, even a large outbreak may be missed entirely, while a smaller but more prolonged outbreak may still permit a detectable effect. In Figure 2, impact is defined by the percent reduction in cases from the number of cases without vaccination to the number of cases with vaccination.

**Figure 2.**
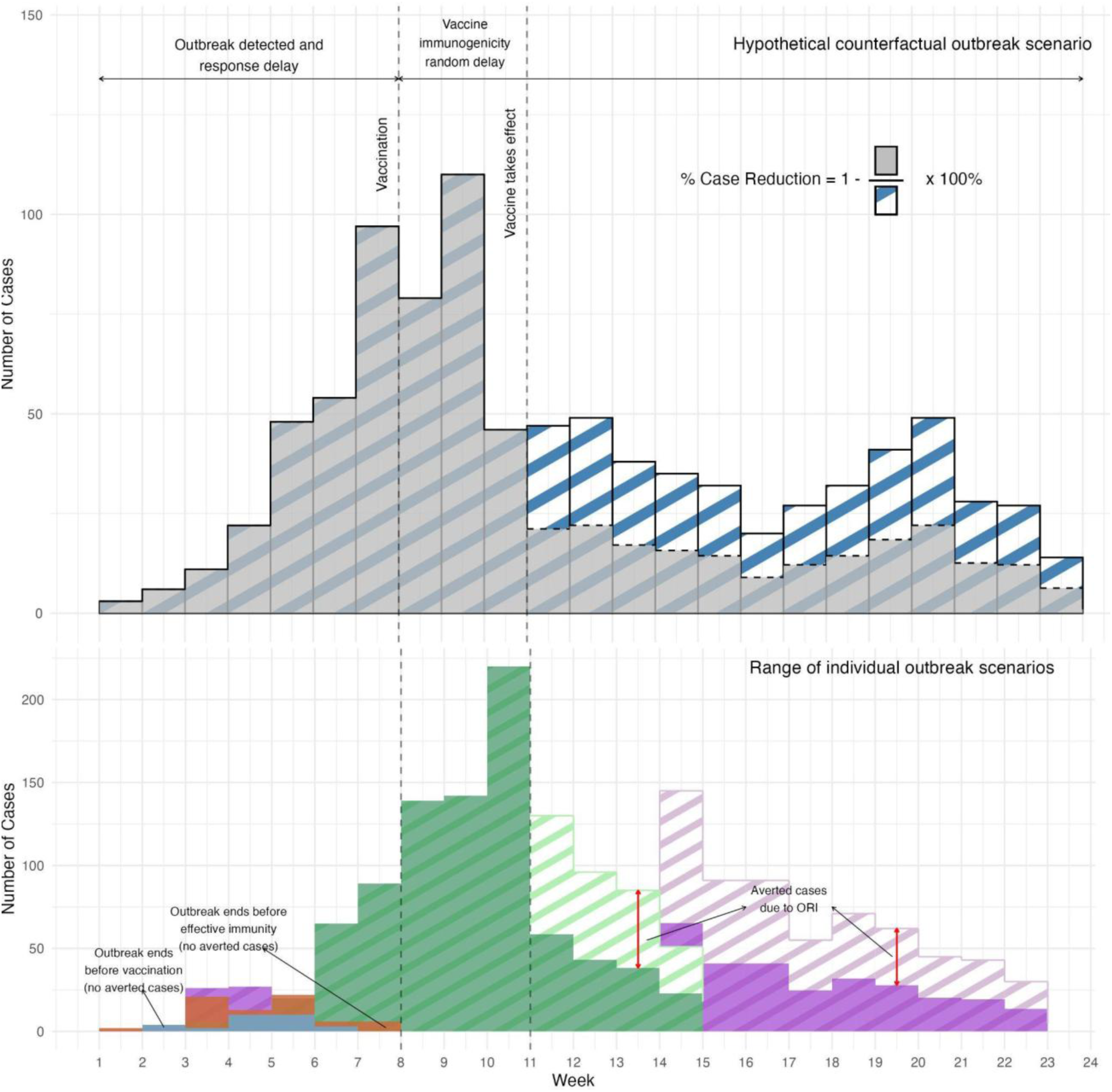
Hypothetical counterfactual outbreak scenarios across the diverse set of individual outbreaks. Blue: Madaya, Myanmar (2000); Orange: Amritsar, India (2019); Green: Harare, Zimbabwe (2011-2012); Purple: Harare, Zimbabwe (2016-2017) Other campaign-, vaccine-, illness-, and cost-related parameters used in the model are found in Table 1. In previous randomized controlled trials of TCVs, seroconversion to vaccine-derived immunity was typically assessed at 28 days following the first dose^17,23–25^. In the study with the greatest number of total participants receiving TCV (n = 283), the rate of seroconversion at day 28 was 98.6% (95% CI: 96.4 – 99.5)^24^. Across all cited studies, the rate of seroconversion at 28 days post-vaccination ranged from 87.8% to 100%. Since this time point was selected to ensure the capture of all immunologic responses, we adopted this as the maximum time frame in our study. However, it is plausible that some individuals may have had prior exposure to *S.*Typhi, resulting in partial or full natural immunity. In such cases, the first dose of TCV may act as a booster, resulting in a more rapid or enhanced immune response^26^. This is further supported by evidence showing that the second dose of TCV produces only a modest increase in antibody titers, likely because levels are already elevated following the initial dose^27^. To account for this possibility, we defined 14 days post-vaccination as the lower bound for seroconversion.

**Table 1.** Model parameters.

| Description | Value | Source |
| --- | --- | --- |
| Campaign-related |  |  |
| Vaccine coverage (%) | 80% (63 – 95) | 30 |
| ORI timing delay (weeks) | 8 | Assumed<br>from data |
| Campaign duration (days) | 14 (3 – 21) | 31 |
| Vaccine-related |  |  |
| Delay in vaccine-derived immunity (days) | 21 (14 – 28) | 17,23–25 |
| Direct vaccine effectiveness by one dose of TCV | 83% (77–87%) | 32 |
| Indirect vaccine effectiveness | 17% | 33 |
| Typhoid fever-related |  |  |
| Proportion of moderate, severe, GI bleed, and intestinal perforation infection | Moderate: 0.35 (0.26 – 0.443)<br>Severe: 0.4775 (0.38 – 0.574)<br>GI Bleed: 0.17 (0.10 – 0.257) | 14 |
|  | Intestinal Perforation: 0.0025 (0 – 0.02) |  |
| Disability weight for moderate, severe, GI bleed, and intestinal perforation infection | Moderate: 0.051 (0.032 – 0.074)<br>Severe: 0.133 (0.088 – 0.19)<br>GI Bleed: 0.325 (0.209 – 0.462)<br>Intestinal Perforation: 0.324 (0.22 – 0.462) | 34 |
| Duration of illness (days) | Moderate: 14 (7 – 21)<br>Severe: 28 (14 – 49) | 35 |
| Burden of AMR cases relative to antimicrobial-sensitive cases | 2 (1 – 3) | 2 |
| Cost-effectiveness |  |  |
| Vaccine cost of procurement per dose | \$1.50 | 36 |
| Vaccine delivery cost per dose | \$0.39 (0.01 – 0.63)** | 2 |
| Vaccine injection and safety equipment per dose | \$0.23 | 2 |
| Cost per outpatient | \$36.58 (0 – 149)** | 2 |
| Cost per inpatient | \$148.76 (20 – 457)** | 2 |
| Life expectancy at mean age of infection | 56 years (46 – 66)*** | 37 |
| Gross Domestic Product (GDP)<br>per capita | \$4,440 (907 – 57,040)*** | 38 |
| Annual percent change average<br>consumer price | From 2017 to 2025: 4.9 (3.2 - 8.6) | 29 |
| Age distribution | Under 2 years: 0.07 (0.06 - 0.08)<br>2 to 4 years: 0.10 (0.09 - 0.11)<br>5 to 16 years: 0.32 (0.31 - 0.35)<br>16+ years: 0.50 (0.45 - 0.52) | 37 |

\*paper reports -0.13, however authors choose to set this to zero since this is not mechanistically possible
\*\*Country-specific, mean (range)
\*\*\*Country- and year-specific, mean (range)

Direct and indirect vaccine effectiveness and campaign duration were sampled (n = 200) from parametric distributions using Sobol sequence methods^28^. Impact and cost-effectiveness of outbreak response immunization could be greater in areas that have positive AMR cases, and thus the burden of AMR was set to twice the burden of antimicrobial-sensitive cases, in line with previous studies^2,8^. All costs were transformed using the global annual percent change in average consumer prices and reported in 2025 USD$^29^. Outbreaks without reported target population size in the original data (n = 7) were supplemented with imputed values obtained from online sources (Supplementary Table 1), corresponding to the smallest administrative level described in the original outbreak report.

Eleven outbreaks reported the implementation of at least one community-based large-scale outbreak response intervention. Among these, water source chlorination was the most common, reported in 7 outbreaks (58%), followed by water, sanitation, and hygiene (WASH) interventions in 5 (42%), public health education campaigns in 4 (33%), antimicrobial administration in 2 (17%), and active surveillance in 1 outbreak (8%). The median time to first intervention was 7 weeks after outbreak onset, with delays ranging from 3 weeks (in an outbreak that implemented chlorination) to a maximum delay of 13 weeks (in an outbreak where the specific intervention type was not reported). This 7-week median timing reflects a realistic benchmark for the mobilization of outbreak responses and was therefore used as the assumed delay in the following analyses. It represents a feasible target timeframe within which vaccination campaigns could be launched, based on observed delays for other similar intervention types. Since the analysis includes monthly outbreaks, this delay was increased to 8 weeks to estimate impact and for comparability.

To assess the robustness of our estimates, sensitivity analyses assessed varying ORI timing, vaccine coverage, and indirect vaccine effectiveness.

### Data availability

All code and processed datasets generated during this study are publicly available in a GitHub repository (https://github.com/kimfinale/typhoid_outbreak_vaccine_impact_modeling/tree/main). The repository contains the complete analysis workflow, including scripts for data cleaning, outbreak reconstruction, statistical modelling, and figure generation, enabling full reproducibility of the results.

## Results

Across the 19 studied outbreaks, a total of 32,573 suspected typhoid cases occurred between years 2000 to 2020, in which 26,298 cases were reported in outbreaks reporting weekly incidence. Figure 3 depicts the monthly times series of each individual outbreak. Fifteen outbreaks occurred in the WHO African region (79%), 2 outbreaks occurred in the South-East Asian region (10.5%), and 2 outbreaks occurred in the Eastern Mediterranean region (10.5%). The median outbreak duration was 23 weeks (ranging from 7 to 143 weeks), median outbreak size was 916 cases (ranging from 33 to 10152 cases) and median attack rate was 1.13 (ranging from 0.6 to 8.1). The median delay for timing of any community-based intervention was 7 weeks (ranging from 2.7 to 11 weeks).

**Figure 3.**
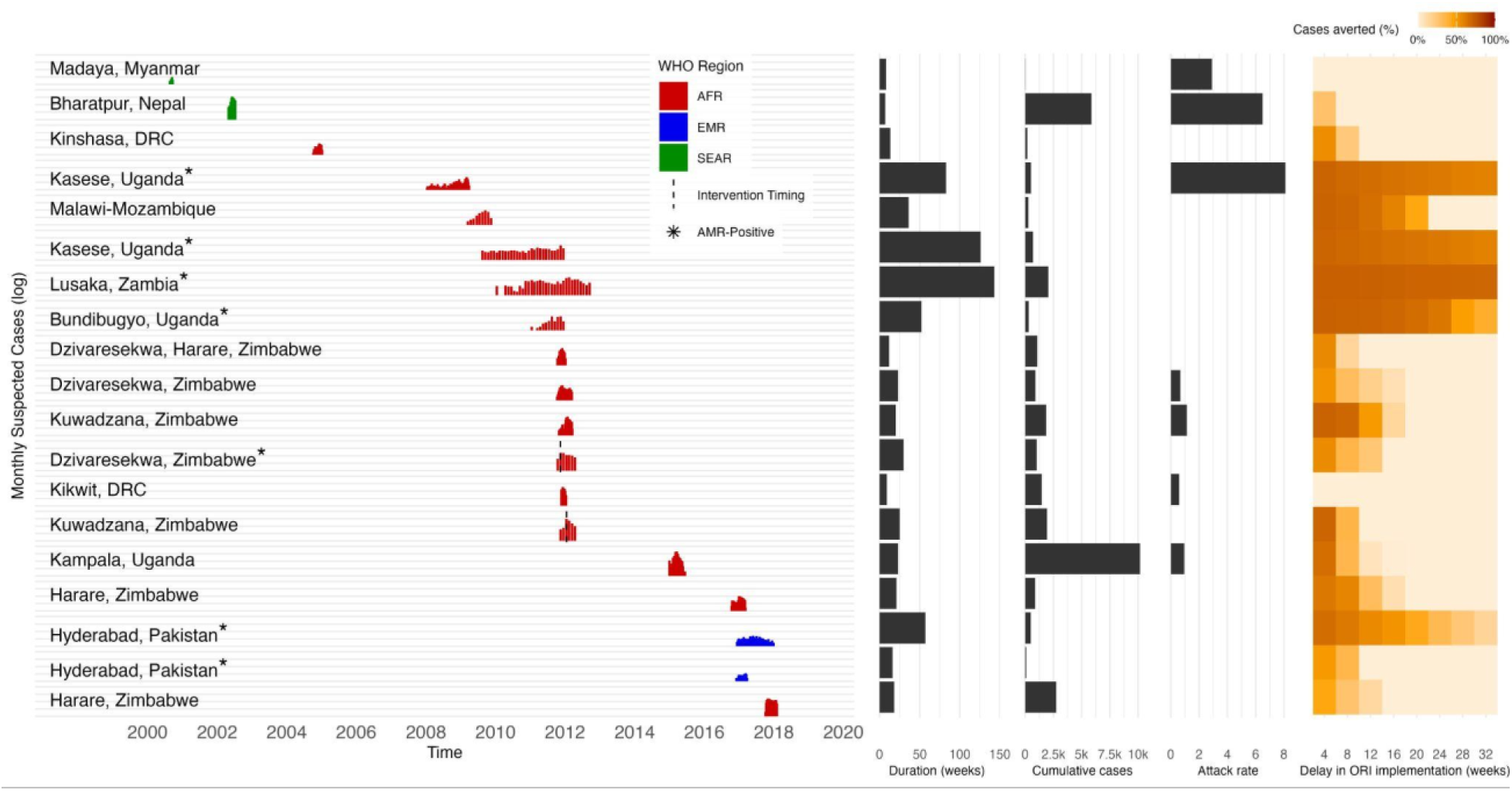
Monthly time series, intervention timing, and AMR-status, and duration, cumulative cases, attack rates, and percent case reduction across delays in ORI (weeks) for 19 outbreaks included in the impact modeling.

Out of the 19 outbreaks, 13 (68%) had at least one AMR-positive case, 1 (5%) outbreaks were negative for AMR, and 5 (26%) outbreaks did not test for AMR. A total of 977 isolates were tested for AMR, in which 889 tested positive. Of those that tested positive, 791 were MDR, 657 were FQ-NS, and 587 were XDR.

In a subset of outbreaks reporting weekly incidence (n = 13), at 80% vaccine coverage, initiating ORI with a 1-week delay led to a median percent case reduction of 66.9% (95% CI: 55.0% – 71.5%), translating to a median of 619 cases averted (95% CI: 32 – 6,129). The median cases averted per 1,000 doses at this delay was 2.5 (95% CI: 0.10 – 42.2) and the median cost per DALY averted was $82,756 (95% CI: 0 - 618,927) (Figure 4). Between week 1 and week 4, the percent case reduction declined by a median of 13.6% (95% CI: 0.9 - 55.9) (or about 4.5% a week), resulting in a 53.9% median case reduction (95% CI: 0% – 70.5%) by week 4. Between week 4 to week 8, the percent case reduction declined by a median of 14.6% (95% CI: 0 - 38.1) (or about 3.7% a week). The percent case reduction reached half of the impact at 1-week delay by week 7 (35.9%, 95% CI: 0% – 68.7%) and week 8 (30.0%, 95% CI: 0% – 68.5%). Similarly, at an 8-week delay in ORI, total cases averted fell to 275 (95% CI: 0 – 2,787), cases averted per 1,000 doses dropped to 0.63 (95% CI: 0.06 – 8.8), and the cost per DALY averted rose to $82,756 (95% CI: 0 – 618,927).

**Figure 4.**
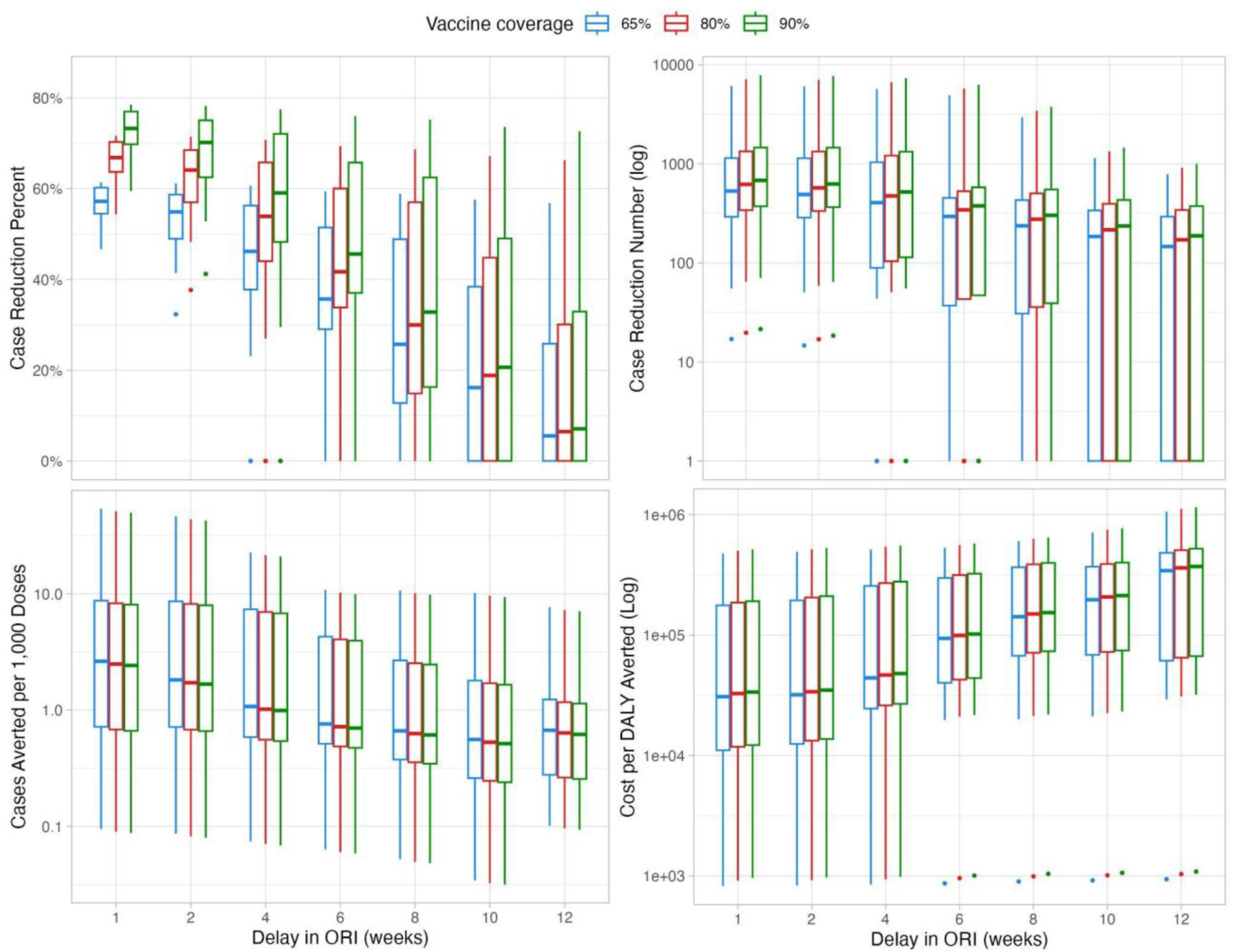
Impact of ORI in weekly reported outbreaks on percent case reduction, case reduction number, cases averted per 1,000 doses, and cost per DALY averted by delay in ORI in weeks and at 65%, 80%, and 90% coverage.

**Figure 5.**
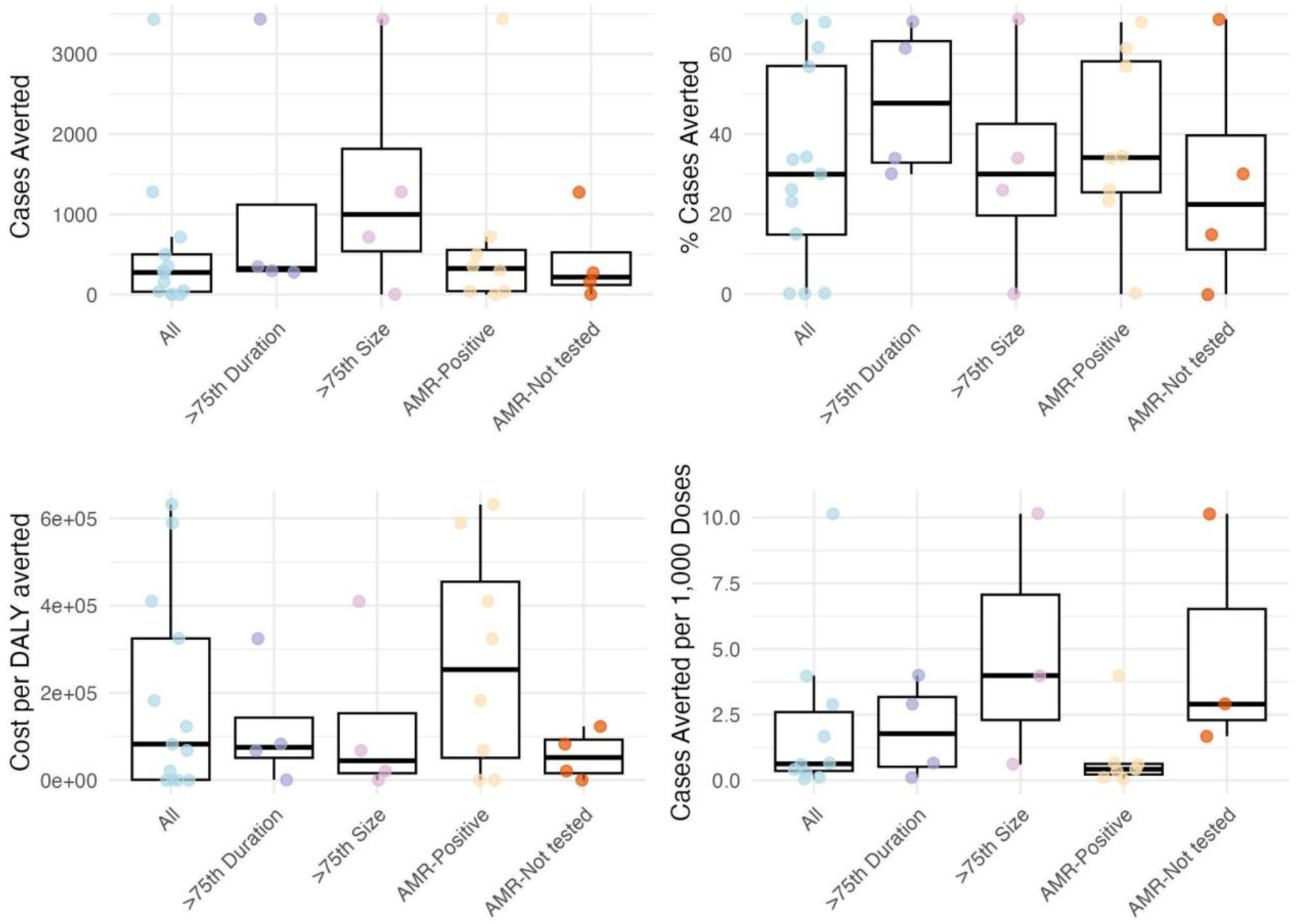
Impact of ORI in all outbreaks reporting weekly incidence and outbreaks stratified by duration, size, and AMR status at 8-week delay and 80% coverage. Median (IQR). Outbreaks that were AMR-Negative (n = 2) had no impact and were excluded from the figure.

As vaccine coverage increased, cases averted as well as the cost per DALY averted also increased, while the cases averted per 1,000 doses decreased. For instance, at 65% coverage, the percent case reduction was 57.2% (95% CI: 47.2% – 61.2%), with 531 cases averted (95% CI: 28 – 5,250), a cost per DALY averted of $25,916 (95% CI: 994 – 249,055), and 2.6 cases averted per 1,000 doses (95% CI: 0.10 – 44.5). At 90% coverage, percent case reduction rose to 60.3%, with 678 cases averted (95% CI: 35 – 6,714), a higher cost per DALY averted of $28,345 (95% CI: 1,144 – 269,865), and 2.4 cases averted per 1,000 doses (95% CI: 0.10 – 41.1).

Outbreaks stratified by duration, size, and AMR status showed substantial variation in the impact of ORI. Outbreaks lasting longer than the 75th percentile tended to have smaller cumulative cases, with a median of 715 cases. Meanwhile, outbreaks with cumulative cases greater than the 75th percentile tended to be of shorter duration, with a median of 19 weeks long. At 80% vaccine coverage and an 8-week delay, outbreaks lasting longer than the 75th percentile (23 weeks) had a higher median number of cases averted (324, 95% CI: 301–348) and greater percent case reduction (64.8%, 95% CI: 61.8%–67.8%) compared to all outbreaks (30.0%, 95% CI: 0% – 68.5%). However, these outbreaks also had higher costs per DALY averted ($162,861, 95% CI: 9,085–316,638) and lower cases averted per 1,000 doses (0.38, 95% CI: 0.12 – 0.64), assuming uniform distribution of vaccines across the administrative unit. In contrast, outbreaks larger than the 75th percentile showed higher median cases averted (719, 95% CI: 36–3,298), more cases averted per 1,000 doses (2.3, 95% CI: 0.79–3.9), and lower cost per DALY averted ($67,866, 95% CI: 3,393– 392,744), but had a lower percent case reduction (26.2%, 95% CI: 1.3–33.4).

AMR-positive outbreaks tended to be both longer (mean: 48 weeks) and larger (mean: 1,945 cumulative cases) than outbreaks that did not test for AMR (mean duration: 16 weeks; mean size: 1,327 cases). These AMR-positive outbreaks had a higher median percent case reduction (34%, 95% CI: 4.1–66.8) and cases averted (324, 95% CI: 6.1–2,959), but also a higher cost per DALY averted ($253,392, 95% CI: 173–624,232) and fewer cases averted per 1,000 doses (0.42, 95% CI: 0.06–3.5), compared to all outbreaks. Similar trends were observed when comparing AMR-positive outbreaks to those without AMR testing, with the exception of cases averted per 1,000 doses, which were notably higher in outbreaks without AMR testing (2.9, 95% CI: 1.7–9.8). The two outbreaks that tested negative for AMR showed no impact at 8-week delays in ORI.

Finally, our sensitivity analyses indicated that outbreaks with monthly incidence data showed patterns very similar to those with weekly incidence, with respect to the influence of vaccine timing and coverage on ORI impact (Supplementary Figure 1). Furthermore, varying the IVE from 0% to 50% had minimal effect on the resulting ORI impact measures (Supplementary Figure 2).

## Discussion

This study highlights the considerable potential of outbreak response immunizations (ORIs) to mitigate the burden of typhoid fever outbreaks, based on the modeling of 19 outbreaks reported between 2000 and 2020. While earlier intervention consistently produced the greatest impact, our findings also demonstrate that even delayed responses, such as those implemented 8 weeks after outbreak onset, can meaningfully reduce case burden, particularly in outbreaks characterized by longer durations or higher cumulative incidence. These results underscore the value of ORIs even in scenarios where logistical or operational constraints delay vaccine deployment.

To date, Zimbabwe remains the only country to have received outbreak response support for typhoid from Gavi, the Vaccine Alliance^19^. Our results suggest that ORI campaigns using typhoid conjugate vaccine (TCV) could serve as an important adjunct to routine immunization programs, which have been introduced in several high-burden countries, including Malawi and Pakistan^39^. Importantly, the effectiveness of ORIs depends on outbreak characteristics. Vaccinating during large outbreaks yielded greater absolute case reductions, whereas vaccinating during prolonged outbreaks resulted in higher relative reductions. This pattern likely reflects the inverse relationship observed in our dataset: larger outbreaks tended to be shorter, and longer outbreaks tended to have fewer total cases. Consequently, in the subset of large outbreaks, vaccination prevented a smaller proportion of cases compared with longer outbreaks, yet the number of cases averted remained substantial in absolute terms. These findings underscore the importance of tailoring outbreak response strategies to specific outbreak dynamics, particularly in resource-limited settings where vaccine allocation must be prioritized efficiently. As such, our findings support greater policy and operational investment in ORI capacity as part of a broader strategy for typhoid control. For instance, although the potential global impact of TCV ORI remains uncertain, our analysis estimates that given a total of 26,298 observed cases (therefore 1,461 per year across 18 years) and a total observed target population of 10,147,833 (therefore 563,769 individuals per year), we could avert 1,409 (96%) and 355 (24%) cases of typhoid each year with a 1-week and 8-week delay in ORI, respectively, and 80% coverage. Across the outbreaks reporting monthly incidence, targeting the total population of 157,950 people annually with an ORI at 8 weeks and 80% coverage would result in 205 (59%) cases averted per year. This is likely a conservative estimate, as it is based solely on outbreaks documented in the published literature and included in this study, and many outbreaks may go unreported.

Previous research, including both modeling and field studies, has consistently demonstrated the effectiveness and cost-effectiveness of TCVs in reducing typhoid fever incidence. Most of these studies, however, have focused on routine immunization strategies. For example, Birger et al. modeled the impact of introducing routine TCV at 9 months of age, supplemented by a catch-up campaign, and predicted that such strategies could avert 46% to 74% of symptomatic cases over ten years in countries eligible for Gavi support^3^. Similarly, dynamic transmission models described by Burrows et al. estimated reductions in symptomatic typhoid ranging from 10% to 46% with routine immunization alone, and up to 90% when paired with catch-up campaigns in optimistic scenarios^4^. Economic evaluations have further supported TCV introduction as a cost-effective, and often cost-saving, intervention in moderate- to high-incidence settings^6,7^. However, this remains one of the few real-world examples of outbreak-focused TCV deployment. To our knowledge, our study is the first to use modeling to specifically evaluate the potential impact of ORI with TCV. By simulating ORIs across a diverse set of historical outbreaks, our findings expand the evidence base to include reactive vaccination scenarios, offering important insights into how TCV could be strategically used beyond routine immunization to mitigate ongoing outbreaks.

Despite its strengths, our study has several limitations. First, we assumed uniform vaccine coverage within administrative units, which may overestimate target populations and underestimate the benefits of more spatially targeted vaccination strategies. The previous Harare outbreak targeted nine suburbs with the greatest impact, rather than all of Harare^31^. Spatially focused TCV delivery, such as targeting known high-incidence areas or outbreak hotspots, may achieve greater impact with fewer doses, which is a nuance our model does not capture. Conversely, our assumption of perfect administrative coverage may overestimate vaccine impact. Reactive and routine TCV campaigns face real-world challenges such as cold chain limitations, expiration or spoilage, which contribute to vaccine wastage. For example, post-campaign evaluations of TCV delivery in outbreak settings have shown that administrative coverage may not reflect true immunization rates for all subgroups^40^. However, in the sole Gavi-approved reactive mass vaccination campaign reported in Zimbabwe, vaccine wastage was reported as less than 0.01%^31^. Although we cannot directly quantify these biases, they may partially offset one another. Second, our parameterization of IVE was informed by estimates from a single cluster-randomized trial^33,41^. Although this study offers valuable evidence, its findings are context-specific and may not be fully generalizable to other settings, population structures, or transmission dynamics. Reliance on a single source to characterize indirect effects introduces uncertainty and may misrepresent the true magnitude of herd protection, which could bias our estimates of vaccine impact. Nevertheless, our sensitivity analyses, which varied IVE values across a plausible range (0–50%), demonstrated that assumptions about indirect protection had minimal influence on the estimated percent reduction in cases (Supplementary Figure 2). This suggests that our conclusions are relatively robust to uncertainty in IVE, although further data from diverse settings would strengthen confidence in these estimates. Third, we assume that vaccine impact is restricted to a single outbreak episode. However, evidence suggests that preventive vaccine interventions may confer preemptive benefits by reducing future outbreak risk in the same or neighboring regions^8^. By not modeling these effects, we may underestimate cumulative impact over time. Lastly, our impact estimates are based on the outbreak dataset used, which is limited to a small subset of outbreaks reported in peer-reviewed journals. This introduces selection bias, as these documented outbreaks may differ systematically from unreported incidents (i.e., often being larger, atypical, or subjected to more rigorous surveillance). As such, our findings may not generalize to more typical or undetected outbreak scenarios, potentially limiting the external validity of our conclusions. However, this is the first study to examine such an extensive typhoid outbreak dataset and thus contributes substantially to the current knowledge gap.

## Conclusion

This study provides important insights into the effectiveness of ORIs across a broad range of typhoid outbreak scenarios. Our findings underscore the importance of timely intervention and strategic outbreak prioritization to maximize vaccine impact. Looking ahead, the results highlight that even with delayed deployment, targeted allocation can substantially reduce cases and improve the overall efficiency of outbreak response. Together, these findings support the integration of ORIs into broader typhoid control strategies, alongside routine immunization and water, sanitation, and hygiene improvements, to mitigate disease burden and enhance epidemic preparedness.

## Data Availability

All data produced are available online at: https://github.com/kimfinale/typhoid_outbreak_vaccine_impact_modeling

## Acknowledgement

This work was supported, in whole or in part, by the Bill & Melinda Gates Foundation [INV-009125]. The conclusions and opinions expressed in this work are those of the authors alone and shall not be attributed to the Foundation.

## Author contributions

Jong-Hoon Kim (J-HK) conceived the study and led the study design and analysis. J-HK and Monica Duong (MD) conducted the statistical analyses and drafted the manuscript. MD curated and prepared the data. MD and J-HK contributed to methodological development and interpretation of results. MD performed the literature review. MD and J-HK contributed to interpretation of findings and refinement of the analytical strategy. MD and J-HK reviewed the manuscript and approved the final version.

## Supplementary

**Supplementary Table 1.** Outbreak-specific target population sizes and sources.

| Outbreak | Target population size | Source |
| --- | --- | --- |
| Michel 2005 | 570 | Original report |
| Aye 2004 | 1102 | Original report |
| Muyembe-Tamfum 2009 | 1648557 | Ferrari et al.<sup>42</sup> |
| Neil 2012 | 670000 | Original report |
| Lutterloh 2012 | 277731 | National Statistical Office of Malawi<sup>43</sup> |
| Hendriksen 2015 | 1747152 | Zambia Statistics Agency<sup>44</sup> |
| Walters 2014, Kasese | 694987 | Uganda Bureau of Statistics<sup>45</sup> |
| Walters 2014, Bundibugyo | 224387 | Uganda Bureau of Statistics<sup>45</sup> |
| Ali 2017 | 400000 | Original report |
| Polonsky 2014, Dzivaresekwa | 118271 | Original report |
| Polonsky 2014, Kuwadzana | 157396 | Original report |
| Muti 2014 | 118271 | Polsonky 2014 |
| Imanishi 2014, Dzivaresekwa | 97000 | Original report |
| Imanishi 2014, Kuwadzana | 129000 | Original report |
| Kabwama 2017 | 1400000 | Original report |
| Qamar 2018 | 3500000 | Original report |
| Davis 2018 | 1485231 | Zimbabwe National<br>Statistics<sup>40</sup> |
| Yousafzai 2019 | 878418 | Original report |
| N'Cho 2019 | 1485231 | Zimbabwe National<br>Statistics<sup>46</sup> |

**Supplementary Table 2.** Impact of ORI in weekly reported outbreaks across 65%, 80%, and 90% coverage. Median (IQR).

| Delay in<br>ORI<br>(weeks) | Vaccine<br>coverag<br>e | Cases<br>averted | % Cases<br>averted | Cost per<br>DALY<br>averted | Cost per<br>case<br>averted | Cost per<br>death<br>averted | Cases<br>averted<br>per 1,000<br>doses |
| --- | --- | --- | --- | --- | --- | --- | --- |
| 1 | 65% | 530.5 (290.8, 1142) | 57.2 (54.5, 60.2) | 30866<br>(14796, 180103) | 1125 (279, 2995) | 0 (0, 21665) | 2.6 (0.7, 8.7) |
| 4 | 65% | 404.7 (88.2, 1036.5) | 46.2 (37.8, 56.3) | 34250 (5836, 229440) | 435 (190, 2971) | 0 (0, 0) | 1.1 (0.6, 7.4) |
| 8 | 65% | 235.6 (29.8, 429.5) | 25.7 (12.8, 48.9) | 78175 (898, 307520) | 1156 (197, 5505) | 0 (0, 0) | 0.7 (0.4, 2.7) |
| 16 | 65% | 0 (0, 73.1) | 0 (0, 8.1) | 0 (0, 268350) | 0 (0, 2645) | 0 (0, 0) | 0.4 (0.1, 0.9) |
| 1 | 80% | 619.1 (339.4, 1332.7) | 66.9 (63.7, 70.3) | 32748<br>(15755, 189898) | 1194 (296, 3160) | 0 (0, 24011) | 2.5 (0.7, 8.3) |
| 4 | 80% | 472.6 (103, 1208.5) | 53.9 (44, 65.8) | 36384 (6291, 242381) | 460 (202, 3134) | 0 (0, 0) | 1 (0.6, 7) |
| 8 | 80% | 274.6 (34.8, 501.2) | 30 (14.9, 57) | 82756 (991, 324731) | 1214 (209, 5808) | 0 (0, 0) | 0.6 (0.4, 2.6) |
| 16 | 80% | 0 (0, 85.1) | 0 (0, 9.4) | 0 (0, 283751) | 0 (0, 2797) | 0 (0, 0) | 0.4 (0.1, 0.8) |
| 1 | 90% | 678.4 (371.9, 1459.9) | 73.3 (69.8, 77) | 33724 (16255, 195086) | 1230 (305, 3244) | 0 (0, 25235) | 2.4 (0.7, 8.1) |
| 4 | 90% | 518.1 (112.8, 1324.7) | 59.1 (48.3, 72.1) | 37493 (6527, 248852) | 473 (209, 3217) | 0 (0, 0) | 1 (0.6, 6.8) |
| 8 | 90% | 300.5 (38.1, 549.2) | 32.8 (16.3, 62.5) | 85150 (1040, 333718) | 1244 (216, 5964) | 0 (0, 0) | 0.6 (0.3, 2.5) |
| 16 | 90% | 0 (0, 93.2) | 0 (0, 10.3) | 0 (0, 291806) | 0 (0, 2876) | 0 (0, 0) | 0.3 (0.1, 0.8) |

**Supplementary Table 3.** Impact of ORI in monthly reported outbreaks across 65%, 80%, and 90% coverage. Median (IQR).

| Delay in ORI (weeks) | Vaccine coverage | Cases averted | % Cases averted | Cost per DALY averted | Cost per case averted | Cost per death averted | Cases averted per 1,000 doses |
| --- | --- | --- | --- | --- | --- | --- | --- |
| 4 | 65% | 445.4 (237.2, 984.6) | 60 (59.5, 60.8) | 67268<br>(13521, 151113) | 1578 (425, 2443) | 0 (0, 0) | 1.3 (1, 6) |
| 8 | 65% | 355.4 (209.4, 539.4) | 58.8 (37.2, 60.3) | 71410<br>(19192, 153364) | 1584 (564, 2489) | 0 (0, 0) | 1.3 (1, 4) |
| 16 | 65% | 157.8 (35.8, 333.1) | 52.6 (12.4, 58) | 65339 (1617, 160727) | 1629 (236, 2880) | 0 (0, 0) | 1 (0.8, 1.2) |
| 32 | 65% | 50.4 (0, 286.1) | 17.2 (0, 46.1) | 3338 (0, 146147) | 487 (0, 2934) | 0 (0, 0) | 0.8 (0.7, 1) |
| 4 | 80% | 521.3 (276.9, 1149.8) | 70 (69.4, 71) | 71138<br>(14381, 159528) | 1679 (456, 2578) | 0 (0, 0) | 1.2 (1, 5.7) |
| 8 | 80% | 415.6 (244.9, 631.9) | 68.6 (43.5, 70.3) | 75419<br>(20546, 161930) | 1684 (602, 2626) | 0 (0, 0) | 1.2 (1, 3.8) |
| 16 | 80% | 184.3 (41.9, 388.6) | 61.4 (14.5, 67.7) | 69121 (1748, 169777) | 1732 (255, 3037) | 0 (0, 0) | 1 (0.8, 1.2) |
| 32 | 80% | 58.9 (0, 333.9) | 20.1 (0, 53.9) | 3604 (0, 154342) | 526 (0, 3103) | 0 (0, 0) | 0.7 (0.7, 1) |
| 4 | 90% | 571.8 (303.3, 1259.9) | 76.7 (76, 77.7) | 73156 (14827, 163919) | 1731 (472, 2648) | 0 (0, 0) | 1.2 (1, 5.6) |
| 8 | 90% | 455.8 (268.5, 693.6) | 75.1 (47.8, 77.1) | 77510 (21259, 166399) | 1736 (622, 2697) | 0 (0, 0) | 1.2 (0.9, 3.7) |
| 16 | 90% | 201.8 (45.9, 425.6) | 67.3 (15.9, 74.1) | 71107 (1815, 174482) | 1787 (265, 3120) | 0 (0, 0) | 0.9 (0.8, 1.1) |
| 32 | 90% | 64.6 (0, 365.8) | 22.1 (0, 59) | 3743 (0, 158574) | 547 (0, 3191) | 0 (0, 0) | 0.7 (0.7, 1) |

**Supplementary Figure 1.**
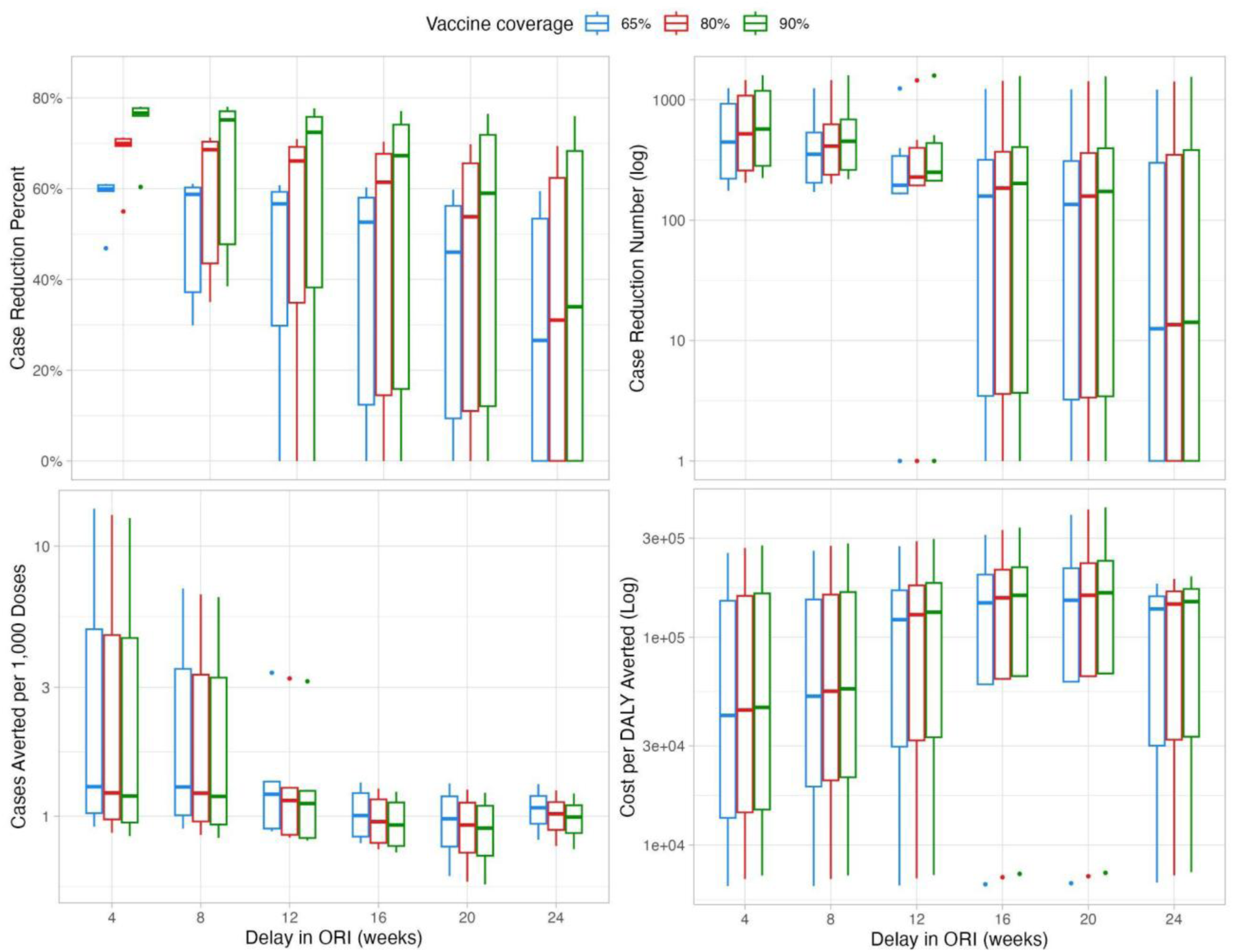
Impact of ORI in monthly reported outbreaks on percent case reduction, case reduction number, cases averted per 1,000 doses, and cost per DALY averted by delay in ORI in weeks and at 65%, 80%, and 90% coverage.

Trends were similar in the subset of outbreaks that reported monthly incidence (n = 6) (Supplementary Figure 1). At an 8-week (or 2-month) delay in ORI and 80% vaccine coverage, median percent case reduction was 69.0% (95%CI: 35.0 - 71.2), corresponding to a median cases averted of 474 (95%CI: 199 - 1,453) cases. Additionally, median cost per DALY averted were $23,770 (95%CI: 6,863 - 275,223) and median cases averted per 1,000 doses were 1.3 (95%CI: 0.9 - 6.6). Impact was affected by vaccine coverage and timing. For example, at an 8-week delay, a 65% vaccine coverage resulted in lower median percent case reduction (59.1%, 95%CI: 29.9 - 61.1) and lower median cases averted (406, 95%CI: 171 - 1,246), and a 90% vaccine coverage resulted in higher median percent case reduction (75.6%, 95%CI: 38.5 - 78.1) and higher median cases averted (519, 95%CI: 218 - 1,593). Furthermore, increasing delay in ORI timing led to generally smaller impact. For example, a 12-week delay resulted in smaller percent case reduction (68.4%, 95%CI: 24.6 - 70.9), cases averted (357, 95%CI: 189 - 1,447), cases averted per 1,000 doses (1.2, 95%CI: 0.8, 3.2), and higher costs per DALY averted ($80,160, 95%CI: 6,909 - 289,662).

**Supplementary Figure 2.**
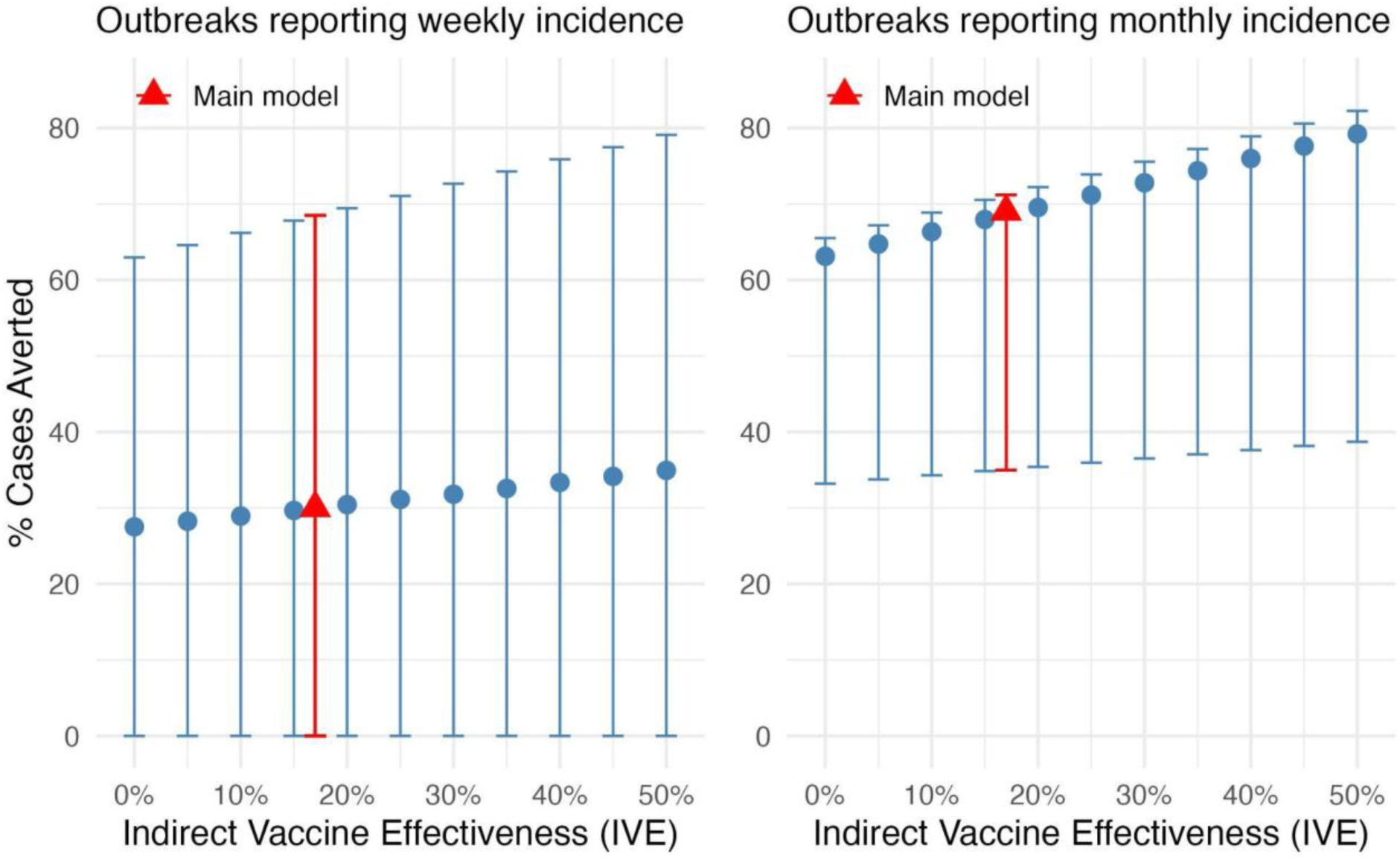
Impact of varying indirect vaccine effectiveness in ORI with 8-week delay and 80% vaccine coverage on percent case reduction (median, 95%CI) in outbreaks reporting weekly and monthly incidence.

Supplementary Figure 2 shows the effect of varying indirect vaccine effectiveness (IVE) on overall median percent case reduction across the subset of weekly and monthly outbreaks. With IVE ranging from 0% to 50% in ORI with 8-week delay and 80% vaccine coverage, the resulting percent case reduction ranged from 27.5% (95%CI: 0 - 63.0) to 34.9% (95%CI: 0 - 79.1) in outbreaks reporting weekly incidence and ranged from 63.1% (95%CI: 33.2 - 65.5) to 79.2% (95%CI: 38.7 - 82.3) in outbreaks reporting monthly incidence. Estimates from the main model are highlighted in red.

